# Level of Physical Activity and ApoE Status - Effects on Alzheimer’s Disease and on Mortality

**DOI:** 10.64898/2026.06.18.26355781

**Authors:** Phillip Ma, Yan Cheng, Yijun Shao, Edward Zamrini, Xuemei Sui, Debby W. Tsuang, Mark Logue, Peter Kokkinos, Ali Ahmed, Charles Faselis, Qing Zeng-Treitler

## Abstract

**Background:** Alzheimer’s disease and related dementias (ADRD) affect over 7.2 million Americans aged 65 and older, with the *APOE* ε4 allele representing the strongest known genetic risk factor. Physical activity (PA) has been associated with reduced dementia risk, but its interaction with *APOE* genotype remains poorly characterized in large, genomically informed cohorts.

**Methods:** We conducted a retrospective cohort analysis using linked genomic, survey, and longitudinal electronic health record data from the VA Million Veteran Program (MVP). Veterans aged ≥65 at enrollment with available *APOE* genotype data, at least 10 years of prior VA medical history, and a completed Lifestyle Survey were included. Individuals with a pre-existing ADRD diagnosis were excluded, yielding a final cohort of 137,593 veterans. Self-reported vigorous physical activity (PA) was ascertained from the MVP Lifestyle Survey and coded as both a four-level ordinal variable and a binary active/inactive classification. The primary composite outcome was incident ADRD or all-cause mortality. Cox proportional hazards models were fitted adjusting for age, sex, race/ethnicity, and baseline comorbidities. A multiplicative interaction term between *APOE* ε4 allele count and PA level was included to formally test for effect modification.

**Results:** Over a median follow-up of 84 months, 65,628 participants (47.7%) experienced the composite outcome. Each additional *APOE* ε4 allele was associated with a 19% increase in risk (aHR = 1.19, 95% CI 1.17–1.21), while active individuals had a 33% lower risk compared to inactive individuals (aHR = 0.67, 95% CI 0.66–0.68). A statistically significant interaction between *APOE* ε4 burden and PA was identified (p < 0.005). Stratified analyses demonstrated that the protective association of PA was present across all genotype groups, with ε4 homozygotes demonstrating a 22% risk reduction among active individuals.

**Conclusions:** In this large veteran cohort, vigorous PA was independently and significantly associated with reduced risk of incident ADRD or death across all *APOE* genotype groups, with the greatest absolute benefit observed among individuals at highest genetic risk. These findings support the prioritization of PA as a targeted preventive strategy, particularly for *APOE* ε4 carriers.

## Introduction

Alzheimer’s disease and related dementias (ADRD) remain among the leading causes of morbidity and mortality in the United States^1^. The Alzheimer’s Association estimated that over 7.2 million people over the age of 65 are diagnosed with ADRD, with incident cases currently near 920,000 per year. The burden extends well beyond those diagnosed, with roughly 12 million Americans providing unpaid care to friends or family members with dementia^2^.

Apolipoprotein E (APOE) is a protein involved in fat metabolism, present in astrocytes and macrophages and found throughout the body. The single gene encoding it has three major alleles: APOE ε2, ε3, and ε4. The ε3 allele is the most common variant (60-75%), with ε4 comprising 15-20%. A single copy of ε4 increases the risk of developing ADRD by a factor of 2-3 relative to non-carriers, while two copies increase that multiplier to between 8 and 12^3-5^. In addition to this increased risk, ε4 is associated with earlier disease onset and faster cognitive decline.

APOE is involved in cholesterol metabolism within the brain as well as in the periphery^4^. Cholesterol is utilized extensively within the brain, being a central part of neuronal growth and repair. It has been shown that higher levels of LDL in serum are associated with cardiovascular disease, stroke, and dementia^6^. APOE ε4 carriers are also known to have higher LDL serum^7^ levels, which could plausibly lead to a higher risk of dementia. Several other studies have shown that exercise has positive effects on serum cholesterol^8^ and have even demonstrated a reduced risk of atherosclerosis specifically in APOE ε4 carriers^9^. Another possible pathway where exercise may interact with ADRD risk is through neuroinflammation. Aβ plaques induce the recruitment of microglia but are supposedly unable to clear the plaques effectively which can lead to cytotoxicity^10^. The involved cytokines include tumor necrosis factor alpha (TNFα), interleukin-1β (IL-1β) and interleukin 6 (IL-6); all of which were significantly increased in mice with APOE ε4^11^. Another animal model study showed that exercise was able to decrease these pro-inflammatory cytokines which led to lower levels of Aβ plaques^12^.

Significant effort and resources have been devoted to developing treatments for Alzheimer’s disease, resulting in the recent approval of lecanemab and donanemab. However, these disease-modifying drugs carry significant risks and have limited effectiveness, which underscores the need to simultaneously pursue effective preventive strategies. For diseases like type 2 diabetes mellitus, lifestyle interventions have proven comparable to some medications in terms of risk reduction, with enduring effects^13^. Prior work by others has provided evidence that lifestyle interventions are also associated with lower risks of ADRD^14,15^.

The U.S. Protect Brain Health Through Lifestyle Intervention to Reduce Risk (US-POINTER)^16^ trial used a structured, two-year multimodal lifestyle intervention which included physical exercise and a healthy diet. The trial demonstrated that this intervention improved cognitive function, including in the APOE ε4 subgroup (n=660). US-POINTER did not, however, individually study the separate components of the intervention and did not use incident ADRD as a specific endpoint. Using data from the Framingham Heart Study, Marino et al. studied a total of 4,324 individuals, tracking their level of physical activity (PA) and finding that higher levels of PA were associated with 41% and 45% lower risk of all-cause dementia at midlife and late life, respectively^14^. This study examined the ε4 allele’s contribution, but due to its limited cohort size, the authors were unable to find statistical significance across many of their estimated effects. A more recent study from the UK Biobank analyzed over 93,000 participants with PA data, genome-wide genotyping, and AD and found that objectively measured PA substantially reduces AD risk regardless of APOE ε4 subgroup^17^.

While the observational relationship between PA, APOE, and ADRD risk has been investigated in other studies, the results have been mixed and limited. Therefore, further investigation is warranted in a larger cohort that combines rich electronic health record (EHR) data, detailed genomics, and survey data. In this study, we aimed to examine the independent and joint associations between PA, APOE, and ADRD in a large population-based cohort from the Veterans across the U.S.

## Methods

### Study Design and Data Sources

This study was a retrospective cohort analysis using data from the VA’s Informatics and Computing Infrastructure (VINCI), which provides longitudinal health records for all inpatient, outpatient, pharmacy, and laboratory data for veterans receiving care at VA healthcare facilities nationwide. This clinical dataset was linked to the VA’s Million Veteran Program (MVP), a large-scale research program providing detailed survey and genomic data from its participants. MVP data were accessed using the Genomic Information System for Integrative Science (GenISIS), MVP’s high-performance computing environment.

The MVP program has already enrolled over 1 million Veterans, including men and women of European, African American, and Hispanic ancestry. Importantly for a study of dementia and aging, nearly half of the MVP cohort is 65 years of age or older and are hence at risk for AD and other forms of dementia. MVP genetic data is linked to electronic health record (EHR) data from the VA Clinical Data Warehouse (CDW), which dates to 1997. Genetically informed ancestry for MVP participants has been determined using the Harmonized Ancestry and Race/Ethnicity (HARE) method. In addition to EHR data, two surveys have been completed by many MVP participants: the Baseline and Lifestyle surveys, available for 625,000+ and 508,000+ MVP participants respectively (MVP v20_1 data).

### Cohort

We identified all MVP participants with available APOE genotype data who were at least 65 years old at the date of their MVP enrollment, which was used as the patient’s index date. To ensure adequate baseline data, we required individuals to have at least 10 years of prior medical history within the VA system. Veterans with a diagnosis of ADRD (based on ICD codes) recorded in the EHR before the index date were excluded. Patients who did not complete the Lifestyle Survey from the MVP were also excluded (**Figure 1**).

**Figure 1.**
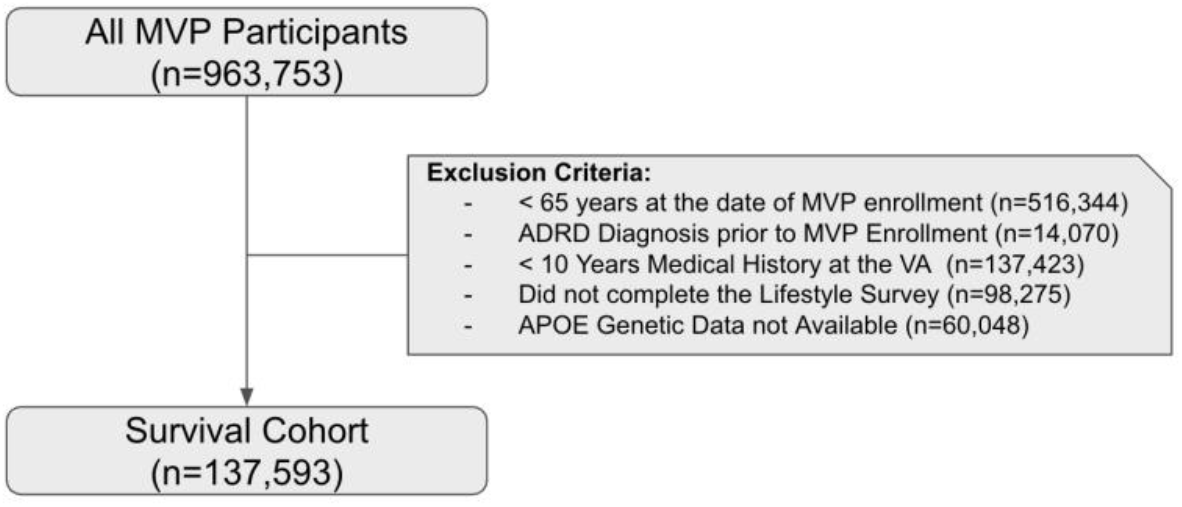
Cohort Creation Diagram

### Exposures

The primary exposures in our analysis were APOE genotype and self-reported PA. APOE genotypes were obtained from pre-populated tables derived from MVP’s genotyping array, and individuals were categorized as ε3/ε3, ε3/ε4, ε4/ε4, ε2/ε3, ε2/ε4, or ε2/ε2. The level of PA was ascertained from the Lifestyle Survey. The response to the question, “How often do you exercise vigorously enough to work up a sweat?” was coded into a four-level ordinal variable: 0 (Rarely/Never), 1 (1-3 times a month or once a week), 2 (2-4 times a week), or 3 (5-6 times a week or daily). To reflect the empirical pattern observed in the survival analyses (described below), a binary active/inactive classification was also derived, collapsing levels 1, 2, and 3 into a single active category, with level 0 (Rarely/Never) serving as the inactive reference.

Genotype data processing and cleaning was performed by an MVP Bioinformatics core and provided to participating MVP study groups. Quality control included checks for sex concordance, advanced batch correction, and assessment for relatedness. The chip design and genotype cleaning pipeline have been described elsewhere^18^. APOE genotype will be based on the TOPmed-imputed data for the two APOE isoform-defining SNPs (rs7412 and rs429358) which were well-imputed in MVP (r2>0.95 for both the European-ancestry and non-European ancestry cohort).

### Covariates

Covariates were selected based on known risk factors for ADRD. Demographic data, including age at the index date, sex, and self-identified race/ethnicity, were obtained from VINCI CDW. Baseline clinical comorbidities were drawn from VINCI CDW using ICD codes. These included coronary artery disease (CAD), depression, diabetes, hypertension (HTN), and hyperlipidemia (HLD), which were identified by contributing clinicians as important comorbid conditions which independently contribute risk to both mortality and ADRD (**Appendix Table 1)**.

### Outcomes

The primary outcome was a composite endpoint of either an incident diagnosis of ADRD or all-cause mortality, whichever occurred first. This approach was chosen to account for death as a competing risk for an ADRD diagnosis. All-cause mortality was determined from death data available in MVP.

We extracted and developed phenotypes related to ADRD based on ICD codes, adapted to the difficulties of identifying subjects with a stringent AD diagnosis based on the limitations of VA EMR data noted above. Given the pattern of ICD code usage in the VA EMR, a large portion of the non-specific codes represent AD cases^19^. Our ICD code algorithm for ADRD includes ICD codes for AD, non-specific dementias, and related dementias. (**Figure 2**). According to our algorithm, cases are included if they have received qualifying codes at least twice. In this schema, The ICD-10 F-codes and ICD-9 “codes specifying dementia in the context of another disorder” codes are classified as “other dementias” and are not by themselves counted towards this total for AD or ADRD, but any corresponding AD or ADRD ICD-9 or 10 codes would be counted.

**Figure 2.**
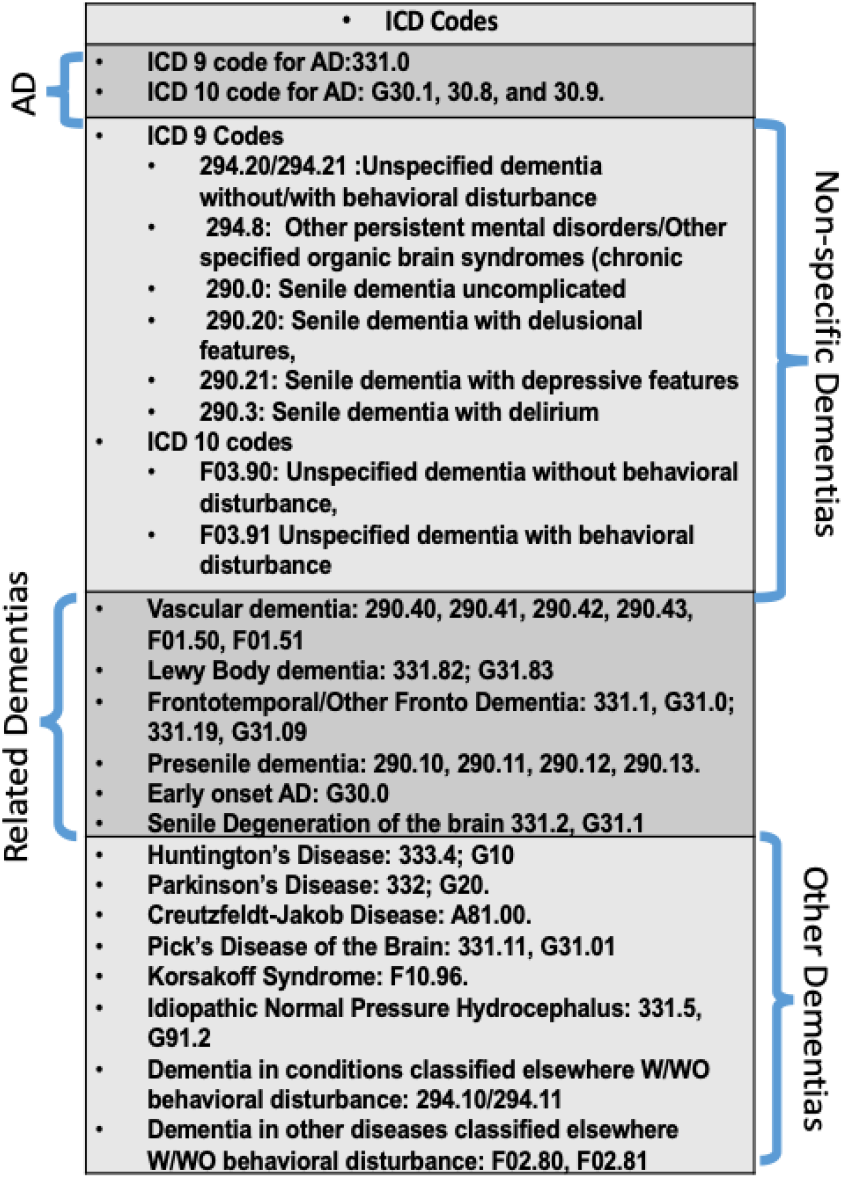
ADRD Phenotyping in MVP

### Statistical Analysis

Baseline characteristics of the cohort were summarized using means, standard deviations, counts, and percentages where applicable. Reported p-values were calculated using t-tests for continuous variables and Chi-square tests for categorical variables.

Kaplan-Meier survival curves were plotted stratified by APOE ε4 allele count and level of activity. Two sets of curves were generated for the activity stratification: one using the four-level ordinal classification and one using the binary active/inactive classification, to assess whether meaningful separation existed across the finer gradations of the ordinal scale. Log-rank tests were used to assess statistical significance between stratified survival curves.

Time-to-event analysis was performed using Cox proportional hazards regression. Proportional hazard assumption was evaluated for each model. Follow-up times were calculated from the index date to the date of the first outcome event (ADRD or death), or to the last visit date in cases where neither event occurred before the study cutoff (June 2025). Four primary Cox models were fitted, all adjusted for age, sex, race/ethnicity, and baseline clinical comorbidities. First, a main-effects model was fitted treating both APOE ε4 allele count and level of activity as ordinal variables to assess their independent associations with the composite outcome. Second, to verify that the ordinal specification did not impose an inappropriate assumption of linearity, both exposures were re-coded as categorical variables and dummy-encoded, with zero ε4 copies and physical inactivity (level 0) serving as the respective reference categories. Third, a model was fitted using the binary active/inactive classification for level of activity while retaining the ordinal specification for APOE ε4 allele count, to capture the clinically relevant activity distinction supported by the Kaplan-Meier analyses. Fourth, an interaction model was fitted including a multiplicative interaction term between APOE ε4 allele count (ordinal) and level of activity (ordinal) to formally test for effect modification.

To illustrate the magnitude of the interaction, predicted 10-year event-free survival probabilities were calculated from the interaction model for representative patient profiles across all 12 permutations of APOE ε4 count and ordinal level of activity. To examine the consistency of the protective association of physical activity across strata of genetic risk, sensitivity analyses were conducted by sub-setting the cohort by APOE ε4 copy number (0, 1, or 2 copies) and fitting separate Cox proportional hazards models within each subset. Each stratified model was run under two specifications: one with level of activity fully dummy-encoded using the inactive category as the reference, and one with the binary active/inactive classification.

The interaction effect between APOE ε4 count and physical activity on the composite outcome was also evaluated. We further conducted sensitivity analyses to stratified sub-cohorts when interaction term was detected significant. Proportional hazard assumptions were also evaluated for each model, by examining log-minus-log plots across level of activity and watching for convergence, the plots for which are provided in **Appendix Figure 1**.

All analyses were conducted using Python 3.10 and the lifelines and scikit-survival packages. A p-value of less than 0.05 was considered statistically significant.

## Results

### Cohort Characteristics

Our final cohort included 137,593 veterans who met all inclusion criteria (**Table 1**). The cohort was predominantly male (97.1%) and White (88.4%), with 8.5% identifying as African American, 0.7% as Asian, and the remaining 2.5% identifying as another race. The mean age was 73.9 years (standard deviation [SD] ± 6.8). Regarding PA, 34.4% of the cohort reported being inactive, with 23.9%, 28.0%, and 13.6% reporting low, moderate, and high levels of activity, respectively. With respect to APOE genotype, 23.4% of the cohort carried at least one ε4 allele, while 1.7% carried two ε4 alleles. Baseline characteristics, including demographics, level of activity, and prevalence of comorbidities, are presented stratified by outcome in Table 1.

**Table 1.** Cohort Characteristics, grouped by Outcome.

| Feature | Value | Overall (%) | No Event (%) | ADRD (%) | Death (%) |
| --- | --- | --- | --- | --- | --- |
| Event Type | Total | 137593 (100%) | 71965 (52.3%) | 16243 (11.81%) | 49385 (35.89%) |
| Age at Enrollment | Mean +/- SD | 73.851 +/- 6.834 | 71.61 +/- 5.272 | 75.851 +/- 7.172 | 76.457 +/- 7.569 |
| Age (Bins) | 65-69 | 46807 (34.02%) | 31251 (43.43%) | 3954 (24.34%) | 11602 (23.49%) |
|  | 70-74 | 37449 (27.22%) | 23066 (32.05%) | 3724 (22.93%) | 10659 (21.58%) |
|  | 75-79 | 23288 (16.93%) | 10581 (14.7%) | 3310 (20.38%) | 9397 (19.03%) |
|  | 80-84 | 17267 (12.55%) | 5164 (7.18%) | 3020 (18.59%) | 9083 (18.39%) |
|  | 85+ | 12782 (9.29%) | 1903 (2.64%) | 2235 (13.76%) | 8644 (17.5%) |
| Gender | M | 133589 (97.09%) | 69500 (96.58%) | 15775 (97.12%) | 48314 (97.83%) |
|  | F | 4004 (2.91%) | 2465 (3.43%) | 468 (2.88%) | 1071 (2.17%) |
| Ethnicity | Not Hispanic Or Latino | 129643 (94.22%) | 67516 (93.82%) | 15187 (93.5%) | 46940 (95.05%) |
|  | Hispanic Or Latino | 7447 (5.41%) | 4225 (5.87%) | 1002 (6.17%) | 2220 (4.5%) |
|  | Unknown | 503 (0.37%) | 224 (0.31%) | 54 (0.33%) | 225 (0.46%) |
| Race | White | 121572 (88.36%) | 63102 (87.68%) | 14122 (86.94%) | 44348 (89.8%) |
|  | Black Or African American | 11672 (8.48%) | 6262 (8.7%) | 1633 (10.05%) | 3777 (7.65%) |
|  | Other | 1874 (1.36%) | 1153 (1.6%) | 229 (1.41%) | 492 (1%) |
|  | American Indian Or Alaska Native | 1081 (0.79%) | 587 (0.82%) | 128 (0.79%) | 366 (0.74%) |
|  | Asian | 947 (0.69%) | 600 (0.83%) | 89 (0.55%) | 258 (0.52%) |
|  | Unknown | 246 (0.18%) | 135 (0.19%) | 26 (0.16%) | 85 (0.17%) |
|  | Native Hawaiian Or Other Pacific Islander | 201 (0.15%) | 126 (0.18%) | 16 (0.1%) | 59 (0.12%) |
| Coronary Artery Disease |  | 55087 (40.04%) | 23163 (32.19%) | 7792 (47.97%) | 24132 (48.87%) |
| Depression |  | 43997 (31.98%) | 22070 (30.67%) | 7410 (45.62%) | 14517 (29.4%) |
| Diabetes |  | 49181 (35.74%) | 23462 (32.6%) | 6488 (39.94%) | 19231 (38.94%) |
| Hypertension |  | 119217 (86.65%) | 59915 (83.26%) | 14801 (91.12%) | 44501 (90.11%) |
| Hyperlipidemia |  | 119209 (86.64%) | 62053 (86.23%) | 14515 (89.36%) | 42641 (86.34%) |
| APOE Genotype | [3, 3] | 84587 (61.48%) | 44553 (61.91%) | 8953 (55.12%) | 31081 (62.94%) |
|  | [3, 4] | 29038 (21.1%) | 14793 (20.56%) | 4489 (27.64%) | 9756 (19.76%) |
|  | [2, 3] | 17543 (12.75%) | 9399 (13.06%) | 1705 (10.5%) | 6439 (13.04%) |
|  | [2, 4] | 3203 (2.33%) | 1679 (2.33%) | 442 (2.72%) | 1082 (2.19%) |
|  | [4, 4] | 2348 (1.71%) | 1081 (1.5%) | 577 (3.55%) | 690 (1.4%) |
|  | [2, 2] | 874 (0.64%) | 460 (0.64%) | 77 (0.47%) | 337 (0.68%) |
| Level of Activity | Inactive | 47391 (34.44%) | 19644 (27.3%) | 6331 (38.98%) | 21416 (43.37%) |
|  | Moderate | 38530 (28%) | 22906 (31.83%) | 4136 (25.46%) | 11488 (23.26%) |
|  | Low | 32925 (23.93%) | 18486 (25.69%) | 3646 (22.45%) | 10793 (21.86%) |
|  | High | 18747 (13.63%) | 10929 (15.19%) | 2130 (13.11%) | 5688 (11.52%) |
| Level of Activity Binary | Active | 90202 (65.56%) | 52321 (72.7%) | 9912 (61.02%) | 27969 (56.64%) |
|  | Inactive | 47391 (34.44%) | 19644 (27.3%) | 6331 (38.98%) | 21416 (43.37%) |
| ADRD |  | 16243 (11.81%) | 0 (0%) | 16243 (100%) | 0 (0%) |
| Months Until Death | Mean +/- SD | 74.755 +/- 37.38 |  | 87.892 +/- 33.446 | 71.898 +/- 37.579 |
| Death After Enrollment |  | 60127 (43.7%) | 0 (0%) | 10742 (66.13%) | 49385 (100%) |

### Survival Analysis

Over a median follow-up of 84.0 months (interquartile range: 58-112 months), a total of 65,628 (47.7%) individuals experienced the primary composite outcome of incident ADRD or Death. 16,243 (11.81%) of participants were diagnosed with ADRD and 49,385 (35.89%) died without having received an ADRD diagnosis. Among those who were diagnosed with ADRD 10,742 (66.13% among that group) had recorded deaths after their diagnosis.

Kaplan-Meier survival curves were plotted to visualize unadjusted event-free survival stratified by APOE ε4 allele count and level of activity. Event-free survival decreased in a stepwise fashion with an increasing number of APOE ε4 alleles, consistent with the well-established dose-response relationship between ε4 burden and ADRD risk (**Figure 3a**). When stratifying by the four-level ordinal activity classification, however, curves for activity levels 1, 2, and 3 did not separate meaningfully from one another; the only appreciable separation was observed between inactive individuals (level 0) and those reporting any degree of activity (levels 1-3) (**Figure 3b**). This pattern suggested that the clinically relevant distinction in this cohort was between physical inactivity and any level of activity, rather than across the finer gradations of the ordinal scale. Accordingly, we also plotted Kaplan-Meier curves using a binary active/inactive classification, which demonstrated clear separation between the two groups (**Figure 3c**).

**Figure 1.**
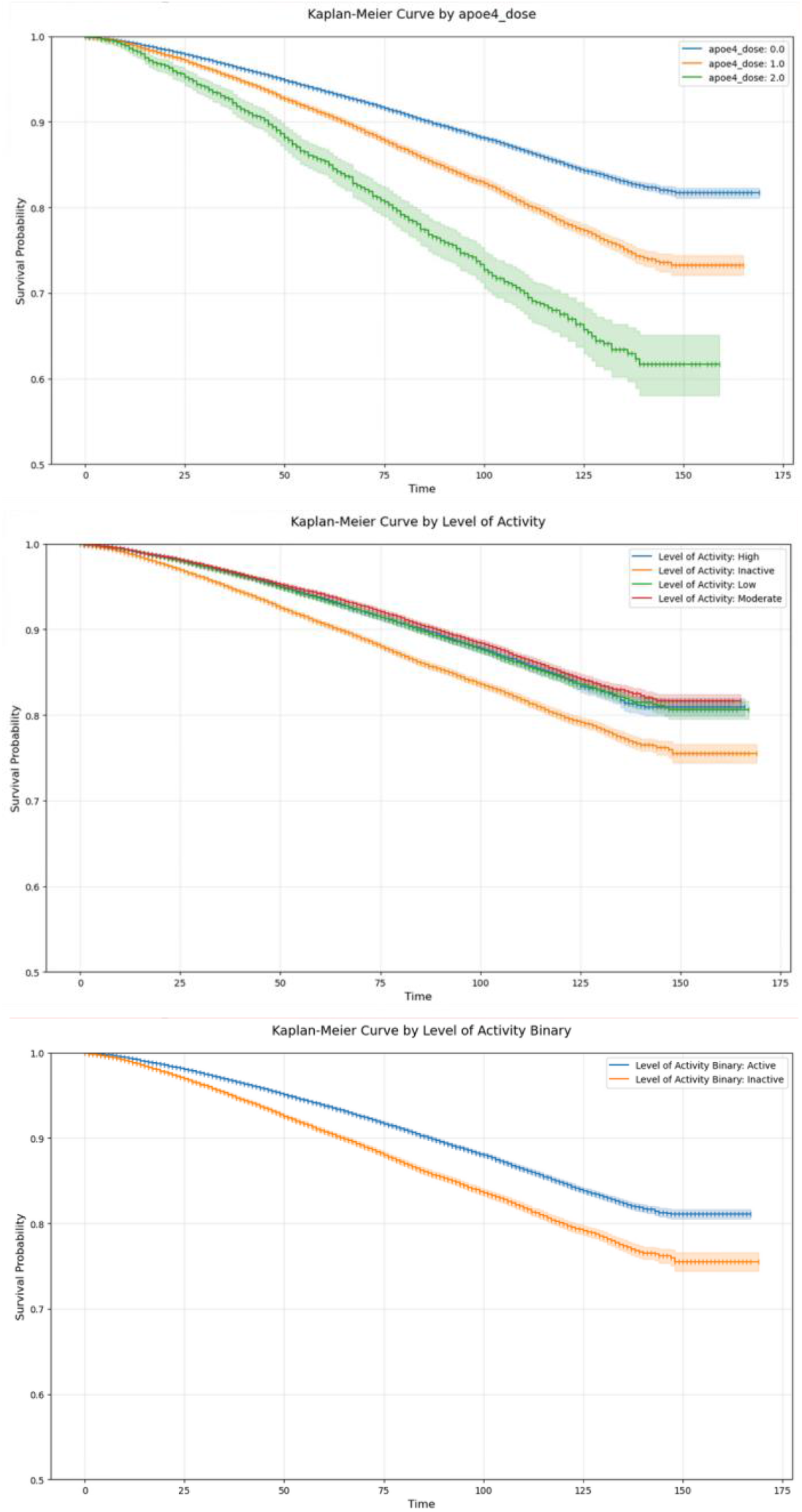
Kaplan-Meier Curves, grouped by **a.** APOE ε4 count, **b**. Level of Activity (ordinal), and **c**. Level of Activity (binary)

In our primary Cox proportional hazards model, adjusted for age, sex, race/ethnicity, and baseline comorbidities, we confirmed the independent effects of both APOE genotype and physical activity (**Table 2**). Both APOE ε4 allele count and level of activity were treated as ordinal variables in this initial specification. Compared to individuals with zero APOE ε4 alleles, each additional ε4 allele was associated with a significantly elevated risk for the composite outcome (aHR = 1.19, 95% CI 1.17-1.21, p < 0.005). Additionally, each one-level increase in self-reported vigorous activity was associated with a meaningful reduction in risk (aHR = 0.84, 95% CI 0.83-0.85, p < 0.005). Additionally, a heat map (**Figure 4**) was produced that shows the mean 10-year survival probability of different subgroups of Level of Activity and APOE ε4 count.

**Table 2.** aHR for ApoE4 Allele Count and Level of Activity Across Variously Encoded Models.

| Model | Variable | aHR |
| --- | --- | --- |
| Main effects (ordinal) | E4 count (per allele) | 1.19 (1.17-1.21)** |
|  | LoA (per level) | 0.84 (0.83-0.85)** |
| Categorical (dummy-encoded) | E4: 1 copy vs. 0 copies | 1.18 (1.16-1.20)** |
|  | E4: 2 copies vs. 0 copies | 1.5 (1.42-1.58)** |
|  | LoA: Level 1 vs. Level 0 | 0.72 (0.71-0.74)** |
|  | LoA: Level 2 vs. Level 0 | 0.64 (0.62-0.65)** |
|  | LoA: Level 3 vs. Level 0 | 0.66 (0.64-0.67)** |
| Binary activity (ordinal E4) | E4 count (per allele) | 1.19 (1.17-1.21)** |
|  | Active vs. Inactive | 0.67 (0.66-0.68)** |

**Figure 4.**
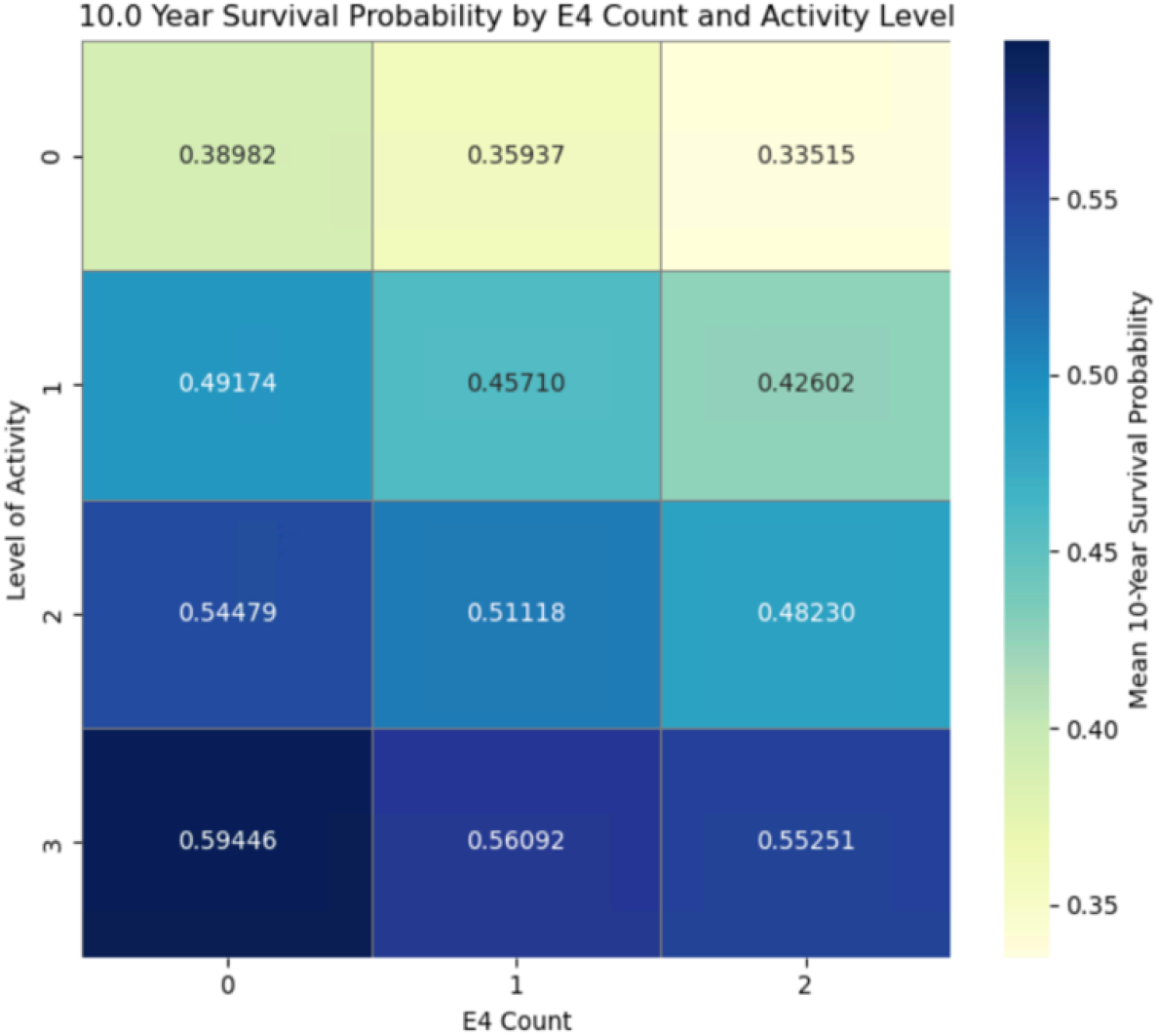
Heat map of Mean 10-Year Survival Probability, grouped by Level of Activity and ApoE4 Count

To verify that treating both exposures as ordinal variables did not impose an inappropriate linearity assumption, we re-ran the primary model with both APOE ε4 count and level of activity fully dummy-encoded, using zero ε4 copies and physical inactivity (level 0) as the respective reference categories (**Table 2**). The results were consistent with the primary model. Relative to inactive individuals, all three active levels were associated with significantly reduced risk (Level 1 vs. 0: aHR = 0.72, 95% CI 0.71-0.74; Level 2 vs. 0: aHR = 0.64, 95% CI 0.62-0.65; Level 3 vs. 0: aHR = 0.66, 95% CI 0.64-0.67; all p < 0.005). For APOE genotype, carrying one ε4 allele was associated with an 18% increase in risk (aHR = 1.18, 95% CI 1.16-1.20, p < 0.005), while carrying two ε4 alleles was associated with a 50% increase in risk (aHR = 1.5, 95% CI 1.42-1.58, p < 0.005). Notably, the hazard ratios across activity levels in the categorical model did not follow a strict pattern, with Level 1 vs Level 0 showing the largest risk reduction. This is consistent with the limited visual separation across the four activity strata observed in the Kaplan-Meier curves and corroborates the decision to also pursue a binary activity specification.

Given this finding, we additionally fitted a model using a binary active/inactive classification for level of activity while retaining the ordinal specification for APOE ε4 count (**Table 2**). Physical activity remained strongly protective and statistically significant under this specification (aHR = 0.67, 95% CI 0.66-0.68, p < 0.005), and the estimate for APOE ε4 count was unchanged (aHR = 1.19, 95% CI 1.17-1.21, p < 0.005).

To formally test for effect modification, an interaction model was fitted that included a multiplicative interaction term between APOE ε4 allele count and ordinal level of activity (**Table 2**). This analysis revealed a statistically significant interaction between APOE ε4 allele count and level of activity (p < 0.005).

### Sensitivity Analysis

To further examine whether the protective association of physical activity was consistent across levels of APOE ε4 burden, we conducted a series of sensitivity analyses by sub-setting the cohort according to ε4 copy number (0, 1, or 2 copies) and fitting separate Cox proportional hazards models (using the composite outcome of ADRD or death) within each subset. Each stratified model was run under two specifications: one with level of activity fully dummy-encoded using the inactive category as the reference, and one with the binary active/inactive classification. Hazard ratios for the activity variables from all stratified models are presented in **Table 3**.

**Table 3.** Sensitivity Analysis.

| **E4 Copies Subset** | **LoA 1 v 0** | **LoA 2 v 0** | **LoA 3 v 0** | **Active (Binary)** |
| --- | --- | --- | --- | --- |
| 0 (n=103,004) | 0.72 (0.7-0.74)** | 0.63 (0.62-0.64)** | 0.64 (0.62-0.66)** | 0.66 (0.65-0.69)** |
| 1 (n=32,241) | 0.72 (0.69-0.75)** | 0.65 (0.62-0.68)** | 0.70 (0.66-0.73)** | 0.69 (0.66-0.71)** |
| 2 (n=2348) | 0.88 (0.76-1.02) | 0.71 (0.62-0.83)** | 0.77 (0.65-0.92)** | 0.78 (0.69-0.88)** |

Among individuals with zero ε4 copies, physical activity was independently significantly protective across all comparisons. Relative to inactive individuals, those at activity levels 1, 2, and 3 had hazard ratios of 0.72, 0.63, and 0.64, respectively (all p < 0.005) after adjusting for other baseline covariates; and the binary active classification yielded an aHR of 0.66 (p < 0.005) with the same adjustment.

Among individuals carrying one ε4 allele, the protective association remained present across all activity comparisons. Adjusted hazard ratios for activity levels 1, 2, and 3 relative to inactive were 0.72, 0.65, and 0.7 (respectively) and each statistically significant (p<0.005). The binary active classification in this subset yielded an aHR of 0.69 (p < 0.005).

Among individuals carrying two ε4 alleles, all but one of the activity comparisons reached statistical significance under either specification (Level 1 vs. 0: aHR = 0.88; Level 2 vs. 0: aHR = 0.71; Level 3 vs. 0: aHR = 0.77; binary active vs. inactive: aHR = 0.78); with the sole non-significant comparison being Level 1 vs 0. This suggests that there is no significant difference between the lowest level of activity and inactivity, though a statistically significant difference was borne out at all levels above and in the binary variant. These findings should be interpreted cautiously and warrant replication in a larger sample of ε4 homozygotes.

## Discussion

### Significance

In this large cohort of U.S. veterans, higher levels of self-reported vigorous physical activity were associated with a reduced risk of the composite outcome of incident ADRD or all-cause mortality. Among ε4 homozygotes, active individuals had 21% reduced risk of developing ADRD or death than inactive individuals, compared to 31% and 34% reduced risk among ε4 one copy carriers and non-carriers, respectively.

Compared to prior studies, the principal strength of our study is the integration of genomic data, lifestyle factors from survey data, and longitudinal EHR data for a very large cohort. This integrated data infrastructure provided the statistical power necessary to detect the nuanced interaction effect.

### Clinical Implications

Our team has previously investigated cardiorespiratory fitness (CRF) as a potential modifiable ADRD risk factor using VA EHR data, finding that higher CRF was associated with lower ADRD risk after controlling for relevant covariates. The present findings extend this line of work by demonstrating that self-reported vigorous physical activity is similarly protective, and that its benefit is meaningfully modulated by APOE ε4 genotype.

These findings invite a reframing of how physical activity is considered in the context of ADRD risk management. Current pharmacological options for Alzheimer’s disease remain limited in their clinical utility. The recently approved anti-amyloid monoclonal antibodies lecanemab and donanemab represent the most significant advances in disease-modifying therapy, yet their benefits are modest, extending functional independence by an estimated 10-13 months^20^, while carrying substantial risks including amyloid-related imaging abnormalities and a considerable cost burden that has been characterized as not cost-effective at current list prices relative to standard of care. Acetylcholinesterase inhibitors and memantine, which remain the mainstay of symptomatic treatment, do not modify the underlying disease course. In this context, physical activity stands out not as a peripheral lifestyle recommendation but as a potentially useful primary prevention tool, one with a favorable risk-benefit profile, low cost, and broad accessibility.

Current guidelines from the Alzheimer’s Association recommend at least 150 minutes of moderate-intensity exercise per week for general brain health, consistent with federal physical activity guidelines. However, these recommendations are largely undifferentiated by genetic risk. Our findings, alongside evidence that APOE ε4 carriers may derive disproportionate cognitive and biomarker benefit from physical activity compared to non-carriers, suggest that genetic risk stratification warrants consideration in how clinicians think about activity counseling. Specifically, identifying individuals as APOE ε4 carriers may provide an opportunity to more deliberately incorporate physical activity into their broader preventive care plan, not as a formal clinical prescription, but as a prioritized and actively reinforced component of risk mitigation conversations. Given the long preclinical window of Alzheimer’s disease, intervening at the level of modifiable behaviors in genetically susceptible individuals before symptom onset represents one of the more practical preventative strategies available to clinicians today.

### Limitations

Nonetheless, our study has important limitations. The primary limitation is our reliance on self-reported physical activity, which is subject to various biases like recall bias. While we plan to incorporate objective CRF data from treadmill tests in future work to validate these findings, it is important to note that activity (a behavior) and fitness (a physiological state) are not synonymous. A complex interplay likely exists between fitness, activity levels, diet, and health outcomes. Therefore, self-reported activity data may continue to provide valuable, complementary information even in models that include objective fitness measures.

A second major limitation is the potential for reverse causation; whereby neurodegenerative changes lead to reduced physical activity rather than the reverse. While our median follow-up of 84 months mitigates this concern to some degree, it cannot be fully excluded. To more formally address this issue, future analyses will employ a lag-time approach, excluding individuals who develop the outcome within the first few years of follow-up to ensure that the lifestyle exposure preceded the disease by a substantial margin. Finally, our cohort consists primarily of male U.S. veterans, which may limit the generalizability of our findings to women and non-veteran populations.

The composite endpoint of ADRD or mortality was prespecified to account for death as a competing risk for ADRD diagnosis; censoring at death in an ADRD-only model would produce biased incidence estimates in a cohort of this age while ignoring it entirely would misattribute deaths to ADRD-free survival. This design follows precedent in aging outcomes trials^21,22^ and ensures that effect estimates reflect the clinically meaningful outcome of remaining both alive and ADRD-free. The principal limitation of this approach is the inability to formally attribute observed effects to either component of the composite endpoint. Formal hazard modeling using the Fine-Gray approach is planned for future work and would provide complimentary estimates to those reported here.

Individuals carrying the APOE ε2 allele were categorized in our genotype exposure variable but were not the focus of the primary analysis. We did not observe enough ε2/ε2 homozygotes to permit a stable stratification and thus focused on APOE ε4. Given the evidence that APOE ε2 is associated with reduced ADRD risk, a complete analysis of APOE allele effects would include them explicitly.

Our findings provide strong observational evidence that physical activity is associated with a lower risk of incident ADRD or death, particularly among individuals at highest genetic risk. While we cannot establish a definitive causal relationship from these data, the consistency of the association and the plausible biological mechanisms are compelling.

## Conclusion

This study leveraged the unique integration of genomic, survey, and longitudinal electronic health record data available in the VA Million Veteran Program to investigate the independent and joint associations between physical activity, APOE ε4 genotype, and the risk of incident ADRD or all-cause mortality in a large cohort of U.S. veterans. Our findings suggest that higher levels of self-reported vigorous physical activity are independently associated with a reduced risk of this composite outcome, and that this protective effect is statistically and clinically meaningful across each of the model specifications that we tested. A significant interaction between APOE ε4 burden and physical activity was identified, with the magnitude of the survival benefit conferred by exercise being greatest among individuals carrying two ε4 alleles. These findings provide strong observational evidence that physical activity may function not only as a broadly protective behavior but also as a means of addressing the elevated risk of ADRD or death conferred by genetics, with the highest-risk individuals standing to benefit the most.

These results carry several limitations: self-reported physical activity may introduce recall bias and does not directly capture cardiorespiratory fitness, and reverse causation (where early neurodegeneration reduces activity) cannot be fully ruled out despite the lengthy follow-up period. The cohort’s composition of predominantly older male veterans limits generalizability to women and non-veteran populations, and the non-significant findings among two-copy ε4 carriers likely reflect insufficient statistical power rather than a true null effect. Future studies should seek to replicate these findings in more diverse cohorts and larger samples of APOE ε4 homozygotes.

Building on the current analysis, several extensions are planned to strengthen the evidence base and address the limitations noted above. To more rigorously address reverse causation, future analyses will employ a lag-time approach, excluding individuals who develop the primary outcome within the first several years of follow-up to ensure that the physical activity exposure substantially preceded disease onset. Objective cardiorespiratory fitness data extracted from VA electronic health records, including performance on treadmill exercise tests, will be incorporated to complement self-reported activity data and provide a more physiologically grounded exposure measure. In parallel, our group is deploying artificial intelligence tools to improve ADRD case ascertainment across the full VA system, which will enhance the precision of outcome definitions and increase statistical power for subgroup analyses. Advanced causal inference methods, including propensity score weighting and doubly robust machine learning approaches, will be applied to reduce confounding and move toward more defensible causal estimates. Together, these efforts will allow for a more granular characterization of the dose-response relationship between vigorous physical activity and ADRD risk, a refinement of effect modification estimates across APOE genotype groups, and ultimately the development of targeted, genetically informed prevention strategies for individuals at highest risk for Alzheimer’s disease and related dementias.

## Data Availability

All data produced in the present study are available upon reasonable request to the authors. All data used is accessible only to researchers approved by the VA.

## Appendix

**Appendix Table 1.**

| Condition | ICD Codes |
| --- | --- |
| Coronary Artery Disease | I25.10, 414.0% |
| Diabetes | E11%, 250% |
| Hypertension | I10, 401.% |
| Hyperlipidemia | E78.01%, E78.2, E78.5, 272.2, 272.4 |
| Depression | F32%, F33%, 296.2%, 296.3% |

**Appendix Figure 1.**
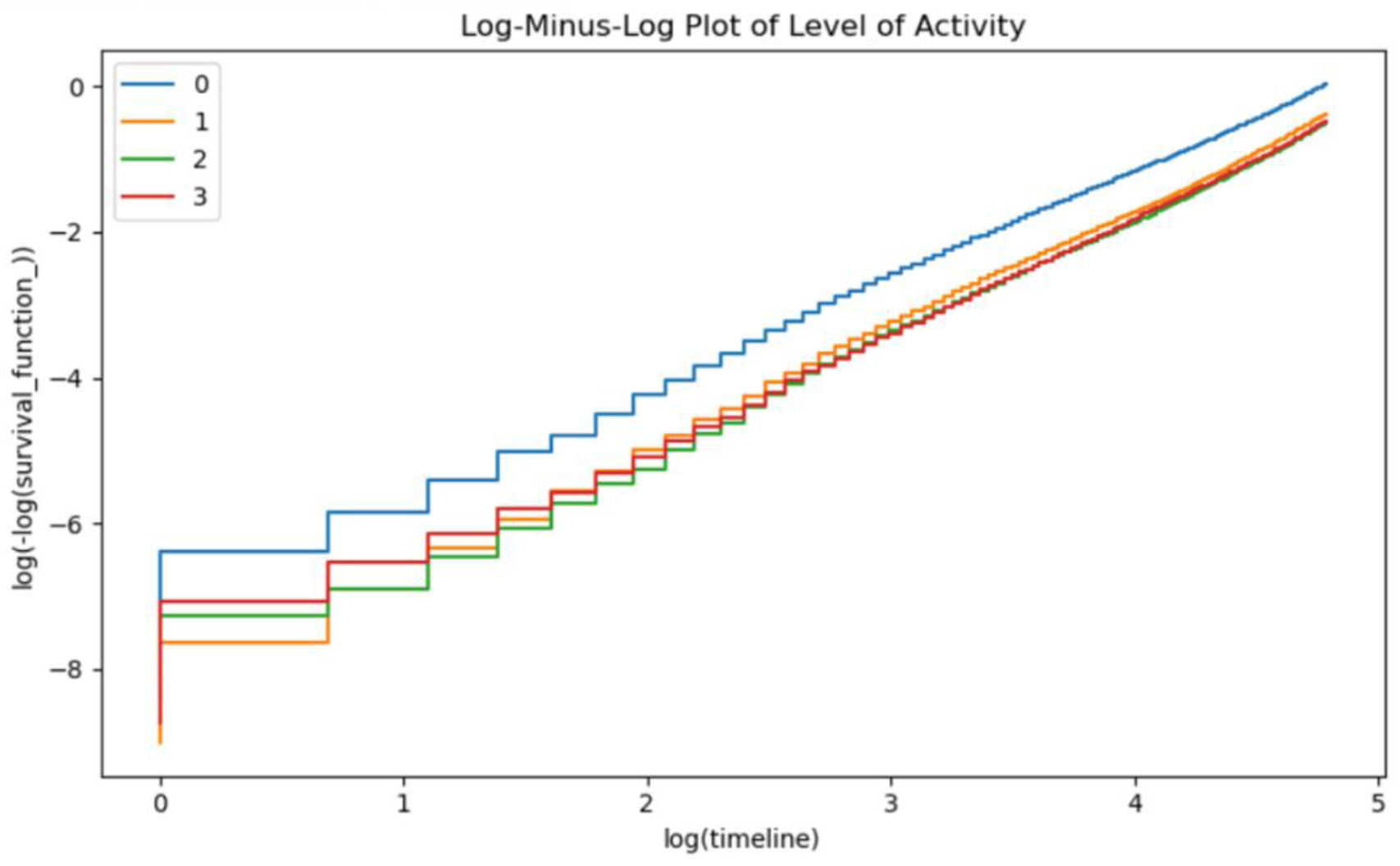

## References

1. 2024 Alzheimer’s disease facts and figures. Alzheimers Dement. May 2024;20(5):3708–3821. doi:10.1002/alz.13809

2. Nandi A, Counts N, Broker J, et al. Cost of care for Alzheimer’s disease and related dementias in the United States: 2016 to 2060. NPJ Aging. Feb 8 2024;10(1):13. doi:10.1038/s41514-024-00136-6

3. Hauser PS, Ryan RO. Impact of apolipoprotein E on Alzheimer’s disease. Curr Alzheimer Res. Oct 2013;10(8):809–17. doi:10.2174/15672050113109990156

4. Liu CC, Liu CC, Kanekiyo T, Xu H, Bu G. Apolipoprotein E and Alzheimer disease: risk, mechanisms and therapy. Nat Rev Neurol. Feb 2013;9(2):106–18. doi:10.1038/nrneurol.2012.263

5. Farrer LA, Cupples LA, Haines JL, et al. Effects of age, sex, and ethnicity on the association between apolipoprotein E genotype and Alzheimer disease. A meta-analysis. APOE and Alzheimer Disease Meta Analysis Consortium. JAMA. Oct 22-29 1997;278(16):1349–56.

6. Lee JH, Hong SM, Shin YA. Effects of exercise training on stroke risk factors, homocysteine concentration, and cognitive function according the APOE genotype in stroke patients. J Exerc Rehabil. Apr 2018;14(2):267–274. doi:10.12965/jer.1836108.054

7. Piccarducci R, Daniele S, Fusi J, et al. Impact of ApoE Polymorphism and Physical Activity on Plasma Antioxidant Capability and Erythrocyte Membranes. Antioxidants (Basel). Nov 9 2019;8(11)doi:10.3390/antiox8110538

8. Brown BM, Peiffer JJ, Taddei K, et al. Physical activity and amyloid-beta plasma and brain levels: results from the Australian Imaging, Biomarkers and Lifestyle Study of Ageing. Mol Psychiatry. Aug 2013;18(8):875–81. doi:10.1038/mp.2012.107

9. Bernstein MS, Costanza MC, James RW, et al. Physical activity may modulate effects of ApoE genotype on lipid profile. Arterioscler Thromb Vasc Biol. Jan 2002;22(1):133–40. doi:10.1161/hq0102.101819

10. Kloske CM, Wilcock DM. The Important Interface Between Apolipoprotein E and Neuroinflammation in Alzheimer’s Disease. Front Immunol. 2020;11:754. doi:10.3389/fimmu.2020.00754

11. Lynch JR, Tang W, Wang H, et al. APOE genotype and an ApoE-mimetic peptide modify the systemic and central nervous system inflammatory response. J Biol Chem. Dec 5 2003;278(49):48529–33. doi:10.1074/jbc.M306923200

12. Corlier F, Hafzalla G, Faskowitz J, et al. Systemic inflammation as a predictor of brain aging: Contributions of physical activity, metabolic risk, and genetic risk. Neuroimage. May 15 2018;172:118–129. doi:10.1016/j.neuroimage.2017.12.027

13. Diabetes Prevention Program Research G, Knowler WC, Fowler SE, et al. 10-year follow-up of diabetes incidence and weight loss in the Diabetes Prevention Program Outcomes Study. Lancet. Nov 14 2009;374(9702):1677–86. doi:10.1016/S0140-6736(09)61457-4

14. Marino FR, Lyu C, Li Y, Liu T, Au R, Hwang PH. Physical Activity Over the Adult Life Course and Risk of Dementia in the Framingham Heart Study. JAMA Netw Open. Nov 3 2025;8(11):e2544439. doi:10.1001/jamanetworkopen.2025.44439

15. Yang JJ, Keohane LM, Pan XF, et al. Association of Healthy Lifestyles With Risk of Alzheimer Disease and Related Dementias in Low-Income Black and White Americans. Neurology. Aug 30 2022;99(9):e944–e953. doi:10.1212/WNL.0000000000200774

16. Baker LD, Espeland MA, Whitmer RA, et al. Structured vs Self-Guided Multidomain Lifestyle Interventions for Global Cognitive Function: The US POINTER Randomized Clinical Trial. JAMA. Aug 26 2025;334(8):681–691. doi:10.1001/jama.2025.12923

17. Na S, Muir K, Lophatananon A, Huang Y, Lee J. Physical activity and Alzheimer’s disease risk across genetic susceptibility: a prospective UK Biobank study using accelerometer data. J Neurol. Dec 2 2025;273(1):1. doi:10.1007/s00415-025-13524-z

18. Hunter-Zinck H, Shi Y, Li M, et al. Genotyping Array Design and Data Quality Control in the Million Veteran Program. Am J Hum Genet. Apr 2 2020;106(4):535–548. doi:10.1016/j.ajhg.2020.03.004

19. Cho K, Gagnon DR, Driver JA, et al. Dementia Coding, Workup, and Treatment in the VA New England Healthcare System. Int J Alzheimers Dis. 2014;2014:821894. doi:10.1155/2014/821894

20. Hartz SM, Schindler SE, Streitz ML, et al. Assessing the clinical meaningfulness of slowing CDR-SB progression with disease-modifying therapies for Alzheimer’s disease. Alzheimers Dement (N Y). Jan-Mar 2025;11(1):e70033. doi:10.1002/trc2.70033

21. Shah RC, Ryan J, Webb KL, et al. Aspirin and healthy lifespan in older people: main outcome of the ASPREE-XT observational study. Lancet Healthy Longev. Sep 2025;6(9):100764. doi:10.1016/j.lanhl.2025.100764

22. McNeil JJ, Woods RL, Nelson MR, et al. Effect of Aspirin on Disability-free Survival in the Healthy Elderly. N Engl J Med. Oct 18 2018;379(16):1499–1508. doi:10.1056/NEJMoa1800722

